# Prevalence of Neurological Soft Signs at Presentation in Pediatric Acute-Onset Neuropsychiatric Syndrome

**DOI:** 10.1101/2024.04.26.24306193

**Authors:** Jane E. Zebrack, Jaynelle Gao, Britta Verhey, Lu Tian, Christopher Stave, Bahare Farhadian, Meiqian Ma, Melissa Silverman, Yuhuan Xie, Paula Tran, Margo Thienemann, Jenny L. Wilson, Jennifer Frankovich

## Abstract

**Importance:** Studies of brain imaging and movements during REM sleep indicate basal ganglia involvement in pediatric acute-onset neuropsychiatric syndrome (PANS). Characterizing neurological findings commonly present in patients with PANS could improve diagnostic accuracy.

**Objective:** To determine the prevalence of neurological soft signs which may reflect basal ganglia dysfunction (NSS-BG) in youth presenting with PANS and whether clinical characteristics of PANS correlate with NSS-BG. Design, Setting, and Participants: 135 new patients who were evaluated at the Stanford Children’s Immune Behavioral Health Clinic between November 1, 2014 and March 1, 2020 and met strict PANS criteria were retrospectively reviewed for study inclusion. 16 patients were excluded because they had no neurological exam within the first three visits and within three months of clinical presentation.

**Main Outcomes and Measures:** The following NSS-BG were recorded from medical record review: 1) glabellar tap reflex, 2) tongue movements, 3) milkmaid’s grip, 4) choreiform movements, 5) spooning, and 6) overflow movements. We included data from prospectively collected symptoms and impairment scales.

**Results:** The study included 119 patients: mean age at PANS onset was 8.2 years, mean age at initial presentation was 10.4 years, 55.5% were male, and 73.9% were non-Hispanic White. At least one NSS-BG was observed in 95/119 patients (79.8%). Patients had 2.1 NSS-BG on average. Patients with 4 or more NSS-BG had higher scores of global impairment (p=0.052) and more symptoms (p=0.008) than patients with 0 NSS-BG. There was no significant difference in age at visit or reported caregiver burden. On Poisson and linear regression, the number of NSS-BG was associated with global impairment (2.857, 95% CI: 0.092-5.622, p=0.045) and the number of symptoms (1.049, 95% CI: 1.018-1.082, p=0.002), but not age or duration of PANS at presentation.

**Conclusions and Relevance:** We found a high prevalence of NSS-BG in patients with PANS and an association between NSS-BG and disease severity that is not attributable to younger age. PANS may have a unique NSS-BG profile, suggesting that targeted neurological exams may support PANS diagnosis.

**Key Points:** *Question:* Do patients with pediatric acute-onset neuropsychiatric syndrome present with neurological soft signs reflective of basal ganglia dysfunction, and are these examination findings associated with disease severity?

*Findings:* In this cohort study of 119 patients with pediatric acute-onset neuropsychiatric syndrome, most patients presented with at least one neurological soft sign pertaining to the basal ganglia. The number of signs was associated with global impairment and the number of PANS symptoms. These findings are consistent with basal ganglia pathology in pediatric acute-onset neuropsychiatric syndrome.

*Meaning:* Targeted neurological exams may help support the diagnosis of pediatric acute-onset neuropsychiatric syndrome.

## Introduction

Pediatric acute-onset neuropsychiatric syndrome (PANS) and pediatric autoimmune neuropsychiatric disorders associated with streptococcal infections (PANDAS) are abrupt-onset neuropsychiatric disorders thought to be triggered by infection.^1,2^ Basal ganglia inflammation may be an important mechanism in these disorders based on imaging studies demonstrating basal ganglia swelling in the acute stage,^3^ microglia activation in the caudate and putamen,^4^ and microstructural changes which are most prominent in the basal ganglia.^5,6^ Additionally, sleep studies in this patient population indicating movements during rapid eye movement (REM) sleep^7–9^ implicate basal ganglia pathology. Parkinson’s disease is a basal ganglia disorder in which movements in REM sleep^10,11^ and the glabellar reflex^12^ can pre-date clinical Parkinson’s disease onset.^13,14^ Interestingly, these findings have also been shown in PANS, suggesting PANS is a basal ganglia disorder.

Autoantibodies targeting cholinergic interneurons in the basal ganglia have been identified,^15^ which could be a cause of imbalance between excitatory and inhibitory synaptic transmission in the basal ganglia and contribute to the cardinal symptoms of PANS (obsessions/compulsions, behavior outbursts, sleep disruption, disinhibition of movements, etc). Moreover, animal models of PANDAS demonstrate an adaptive immune response, involving autoantibodies and Th17 cells, leading to central nervous system pathology including neurovascular injury, blood-brain barrier disruption, activation of microglia, and loss of excitatory synaptic proteins.^16–18^

PANS classification criteria requires abrupt-onset or abrupt-recurrence of obsessive–compulsive symptoms and/or eating restriction with two or more additional new and abrupt-onset neuropsychiatric symptoms which commonly include: emotional lability; irritability, aggression, severely oppositional behaviors; behavioral regression and/or behavior outbursts, deterioration in school performance (often due to new-onset reading and math challenges), sensory amplification, motor abnormalities (most commonly tics, “piano-playing” finger movements, handwriting deterioration, clumsiness), sleep disturbances, and urinary issues (enuresis and urinary frequency).^19–21^

While “hard” neurological exam findings of basal ganglia dysfunction such as chorea or dystonia suggest a condition other than PANS, “neurological soft signs” (NSS) such as voluntary movement overflow have been described in attention-deficit hyperactivity disorder (ADHD),^22–32^ obsessive compulsive disorder (OCD),^22,23,33–38^ autism,^22,23,32,39–43^ and Sydenham chorea,^44–47^ and may also indicate basal ganglia dysfunction. The prevalence of these findings in PANS is not known.

The goal of this study was to characterize the prevalence and predictors of neurological soft signs which may reflect basal ganglia dysfunction (NSS-BG) at clinical presentation. We hypothesized a high prevalence of NSS-BG among patients with PANS and that patients who presented with more NSS-BG would exhibit more severe disease states and experience more PANS symptoms.

## Methods

### Study Setting

This was a retrospective study conducted at the Stanford Children’s Immune Behavioral Health (IBH) Clinic, a multidisciplinary clinic where children and young adults with PANS and PANDAS are followed by multiple sub-specialties including child psychiatry, rheumatology, immunology, pediatrics, and psychology. Informed written consent from parents and adult participants and written assent from competent minor participants were obtained after the nature of the research was fully described and before any data were collected. Data was stored in a secure database. Approval was given by the Stanford University Institutional Review Board (IRB#26922).

### Study Population

The IBH Clinic saw 135 new patients between November 1, 2014 and March 1, 2020 that consented to research and were diagnosed with PANS (eAppendix 1 in the Supplement) by a child psychiatrist (MT, MS, PT, YX). Electronic medical records for the patients in the cohort were retrospectively reviewed. Neurological exams were completed as part of the physical exam by clinicians in the IBH clinic (JF, BF, MM). We included the neurological exam findings from patients’ initial presentation to the IBH clinic. Occasionally, there was no comprehensive neurological exam including NSS-BG done at the first visit, which occurred for various reasons including time restraints, patient distress, or challenges with cooperation. In these cases, the neurological exam from the second or third visit was used for data collection. Patients were excluded if they had no comprehensive neurological exam including NSS-BG within the first three visits and within three months of the first clinic visit (N=16). The final study sample included 119 patients. Most neurological exams included (N=93/119, 78.2%) were from the patient’s first visit to the IBH clinic.

### Sources of Data

Demographics, neurological exam findings, PANS symptoms, and impairment scale scores were abstracted from the electronic medical records and the prospective parent/caregiver questionnaires. These data were maintained within our IRB-approved Immune Behavioral Health REDCap database.

### Measures/Variables

To evaluate exam findings reflective of basal ganglia dysfunction, our clinical protocol was established through a training with a pediatric movement disorder specialist (Dr. Terence Sanger) and several trainings by Dr. Susan Swedo based on her observations of the National Institutes of Health (NIH) PANDAS cohort.^48,49^ All practitioners routinely performed the same exam maneuvers for every patient. Table 1 indicates the procedures and findings for each neurological test used to evaluate patients in our clinic and for this study. These maneuvers were used because they are more likely to reflect basal ganglia dysfunction compared to other neurological tests.

**Table 1.**
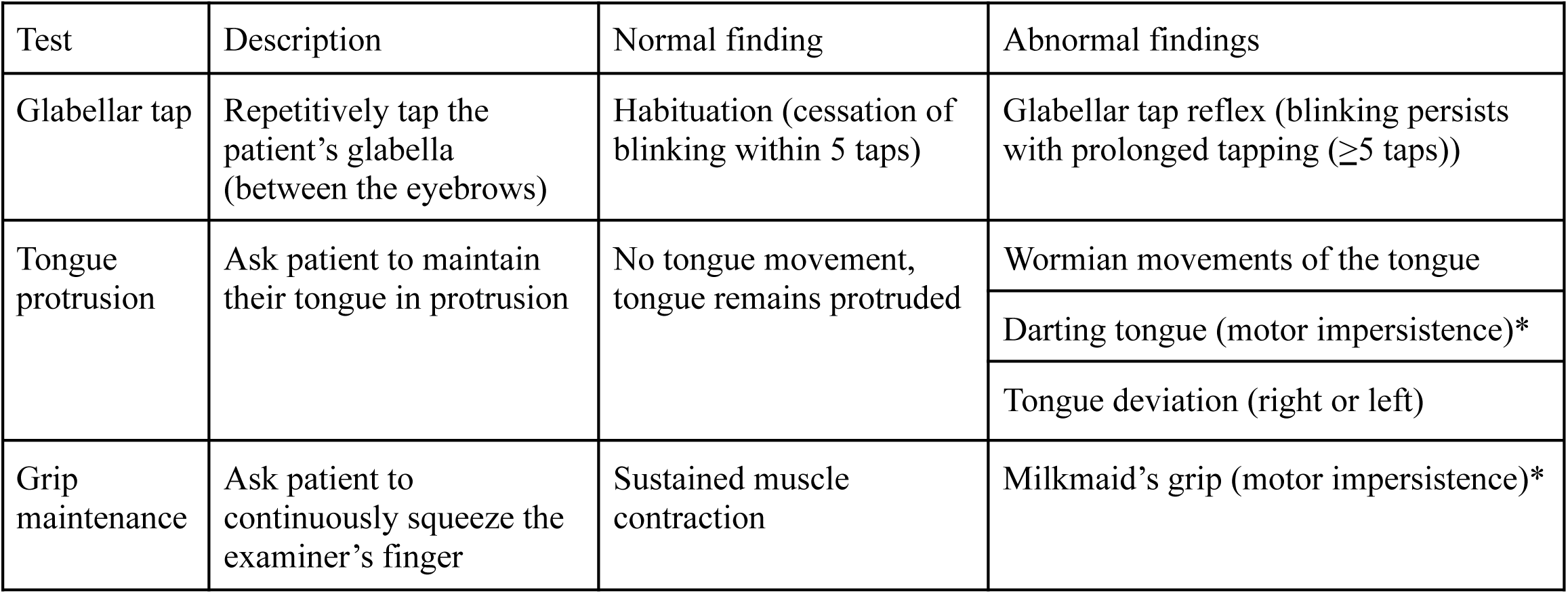
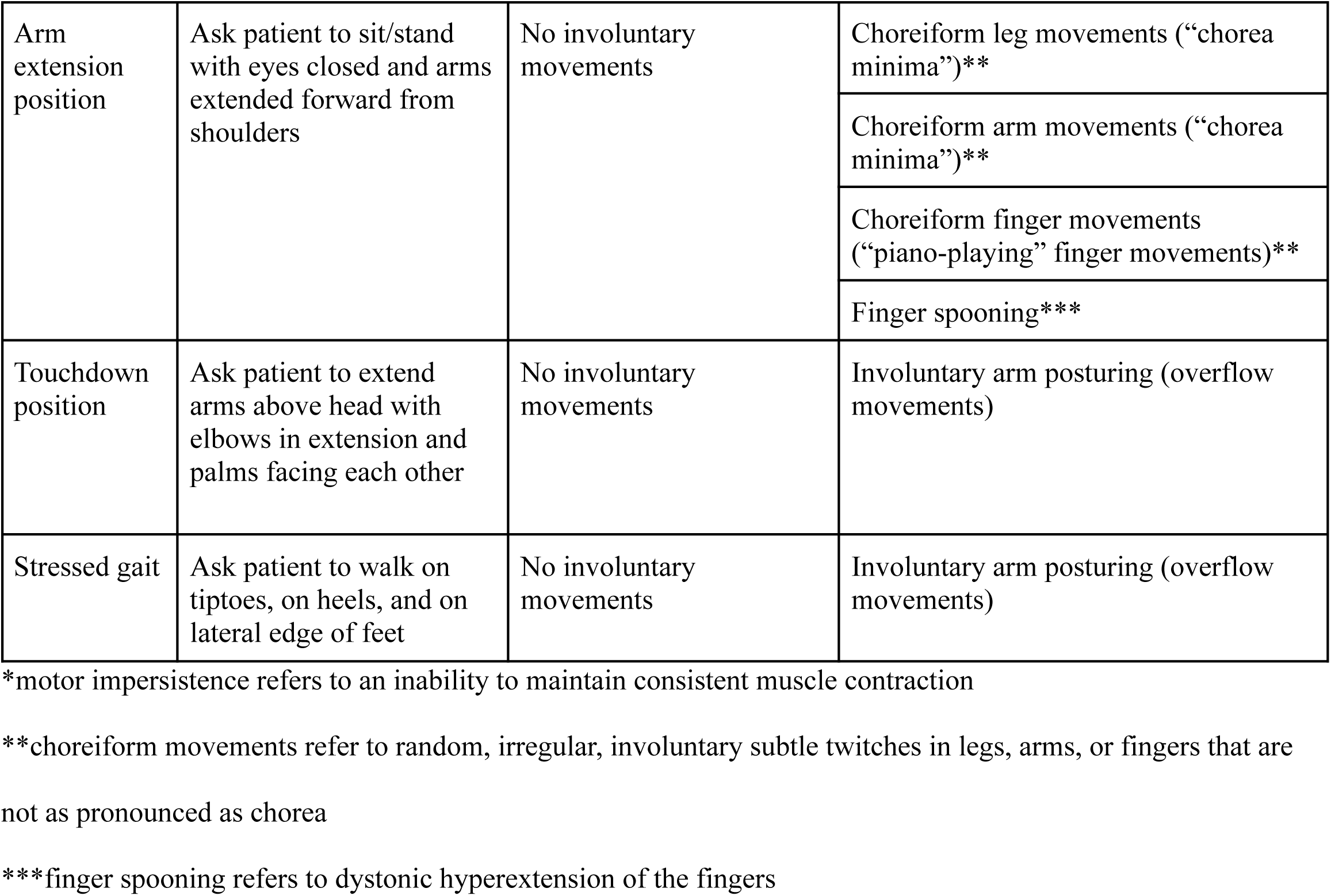
Maneuvers used to evaluate neurological soft signs reflective of basal ganglia dysfunction (NSS-BG) in pediatric acute-onset neuropsychiatric syndrome (PANS)

| Test | Description | Normal finding | Abnormal findings |
| --- | --- | --- | --- |
| Glabellar tap | Repetitively tap the patient's glabella (between the eyebrows) | Habituation (cessation of blinking within 5 taps) | Glabellar tap reflex (blinking persists with prolonged tapping ($\geq 5$ taps)) |
| Tongue protrusion | Ask patient to maintain their tongue in protrusion | No tongue movement, tongue remains protruded | Wormian movements of the tongue |
|  |  |  | Darting tongue (motor impersistence)* |
|  |  |  | Tongue deviation (right or left) |
| Grip maintenance | Ask patient to continuously squeeze the examiner's finger | Sustained muscle contraction | Milkmaid's grip (motor impersistence)* |
| Arm extension position | Ask patient to sit/stand with eyes closed and arms extended forward from shoulders | No involuntary movements | Choreiform leg movements (“chorea minima”)** |
|  |  |  | Choreiform arm movements (“chorea minima”)** |
|  |  |  | Choreiform finger movements (“piano-playing” finger movements)** |
|  |  |  | Finger spooning*** |
| Touchdown position | Ask patient to extend arms above head with elbows in extension and palms facing each other | No involuntary movements | Involuntary arm posturing (overflow movements) |
| Stressed gait | Ask patient to walk on tiptoes, on heels, and on lateral edge of feet | No involuntary movements | Involuntary arm posturing (overflow movements) |

\*motor impersistence refers to an inability to maintain consistent muscle contraction
\*\*choreiform movements refer to random, irregular, involuntary subtle twitches in legs, arms, or fingers that are not as pronounced as chorea
\*\*\*finger spooning refers to dystonic hyperextension of the fingers

The NSS-BG evaluated included: 1) glabellar tap reflex, 2) abnormal tongue movements, 3) milkmaid’s grip, 4) choreiform movements, 5) spooning, and 6) overflow movements. Thus, a patient could present with a minimum of zero or a maximum of six NSS-BG.

Prior to each clinic visit, parents and patients completed an electronic questionnaire including impairment rating scales and questions about the patient’s interim medical and psychiatric symptoms and treatments. The impairment scales used include the Global Impairment Score (GIS), a validated measure of disease severity developed for PANS,^50^ and the Caregiver Burden Inventory (CBI), a measure of caregiver burden that has been validated for patients with PANS.^51–53^ The Global Impairment Score ranges 1-100, and the Caregiver Burden Inventory ranges 0-96; higher scores indicate greater impairment and caregiver burden, respectively.

### Statistical Analysis

Summary statistics were used to describe the demographic and clinical characteristics of our study cohort. Continuous variables were summarized using mean and standard deviation, and categorical variables were summarized using counts and proportions. Selected demographic and clinical characteristics were compared between patients with different NSS-BG prevalences using a two-sample t-test. The following regression models were conducted, with Models 1, 2, and 3 being Poisson regression and Model 4 being linear regression.

● Model 1: Controlling for sex and race, is age associated with the number of NSS-BG?
● Model 2: Controlling for sex, race, and age, is PANS duration associated with the number of NSS-BG?
● Model 3: Controlling for sex, race, and age, is the number of NSS-BG associated with the number of PANS symptoms?
● Model 4: Controlling for sex, race, and age, is the number of NSS-BG associated with the Global Impairment Score? (The analysis was based on 107 patients after excluding 12 patients with missing Global Impairment Score.)

We evaluated confounders of some NSS-BG that could reveal an alternative explanation for the finding, such as age or comorbid conditions. Using a Chi-square test, we conducted two subset analyses: 1) whether or not overflow movements were associated with younger age and 2) whether or not spooning was associated with hypermobility.

All statistical tests were considered to be statistically significant if the two-sided p<0.05. Statistical analysis system (SAS) University Edition (SAS Institute, Inc., Cary, NC, USA) was used for statistical analysis.

## Results

The study cohort included 119 patients with PANS (Table 2). Patients were 3.9-22.4 years of age at presentation (mean=10.4, SD=3.6). The average age of PANS onset was 8.2 years (range=17.0, SD=3.6). Patients presented to the Immune Behavioral Health Clinic an average of 2.2 years (range=11.3, SD=2.7) after initial PANS onset. Most patients were in a flare at the time of evaluation (N=105/119, 88.2%). The cohort was predominantly male (N=66/119, 55.5%) and non-Hispanic White (N=88/119, 73.9%). Obsessions, compulsions, anxiety, sleep disturbance, and irritability were the most common presenting PANS symptoms. Of 29 symptoms assessed via questionnaire, patients presented with 13.2 symptoms on average.

**Table 2.** Demographic and clinical characteristics of 119 consecutive patients with pediatric acute-onset neuropsychiatric syndrome (PANS)

|  | Patients with PANS (N=119) |
| --- | --- |
| Age (years) at PANS onset, mean (SD) | 8.2 (3.6) |
| Age (years) at the first clinic visit, mean (SD) | 10.4 (3.6) |
| PANS duration (years), mean (SD) | 2.2 (2.7) |
| In PANS flare at time of evaluation, N (%) | 105 (88.2) |
| Male gender, N (%) | 66 (55.5) |
| Race and ethnicity, N (%) |  |
| Non-Hispanic White | 88 (73.9) |
| Multiracial | 14 (11.8) |
| White/Other | 13 (10.9) |
| Asian | 3 (2.5) |
| Unknown/Unreported | 1 (0.8) |
| PANS symptoms at clinical presentation, N (%) |  |
| Obsessions | 105 (88.2) |
| Compulsions | 100 (84.0) |
| Hoarding | 18 (15.1) |
| Food refusal/avoidance | 70 (58.8) |
| Urge to overeat | 9 (7.5) |
| Fluid refusal | 13 (10.9) |
| Separation anxiety | 71 (59.7) |
| Other anxiety/fears/phobias/panic attacks | 101 (84.9) |
| Emotional lability | 44 (37.0) |
| Depression/sadness | 75 (63.0) |
| Suicidal ideation/behavior | 19 (16.0) |
| Irritability | 84 (70.6) |
| Aggressive behaviors, violence, and/or rage | 68 (57.1) |
| Oppositional behaviors | 51 (42.8) |
| Hyperactivity or impulsivity | 42 (35.3) |
| Trouble paying attention | 67 (56.3) |
| Baby talk | 10 (8.4) |
| Behavioral/developmental regression | 58 (48.7) |
| Worsening of school performance | 62 (52.1) |
| Worsening of handwriting/copying/art | 31 (26.0) |
| Cognitive impairment | 70 (58.8) |
| Pain (headaches, abdominal pain, body pain) | 78 (65.5) |
| Sensory dysregulation/amplification | 77 (64.7) |
| Motor tics | 43 (36.1) |
| Phonic tics | 25 (21.0) |
| Daytime wetting or bedwetting (enuresis) | 30 (25.2) |
| Urinary frequency (uses restroom frequently) | 37 (31.1) |
| Sleep disturbance | 85 (71.4) |
| Hallucinations | 20 (16.8) |
| Number of above PANS symptoms, mean (SD) | 13.2 (4.3) |
| Impairment scale scores at clinical presentation, mean (SD) |  |
| Global Impairment Score (GIS)<sup>a</sup> | 43.2 (23.2) |
| Caregiver Burden Inventory (CBI)<sup>b</sup> | 32.1 (17.1) |
| Hypermobility at clinical presentation, N (%) | 14 (11.8) |

<sup>a</sup>12 patients (10%) had missing GIS.
<sup>b</sup>44 patients (37%) had missing CBI.

NSS-BG were common in our cohort with 59.7% of subjects presenting with overflow movements, 52.1% with choreiform movements, 31.1% with abnormal tongue movements, 30.2% with milkmaid’s grip, 21.8% with glabellar tap reflex, and 17.6% with spooning (Table 3). These proportions were notably higher among the subset of patients who had a complete neurological exam, meaning they were examined for all six NSS-BG.

**Table 3.**
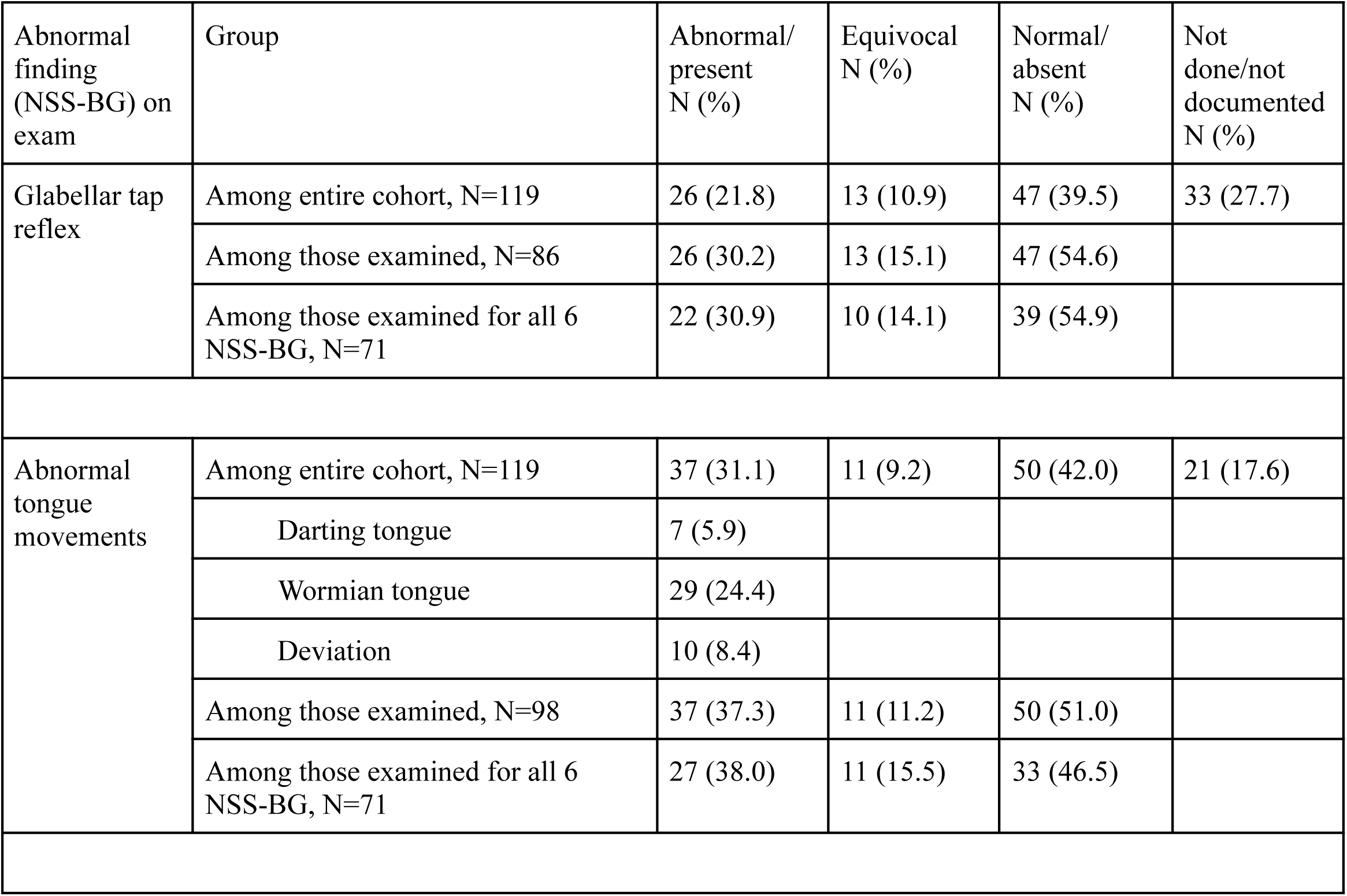
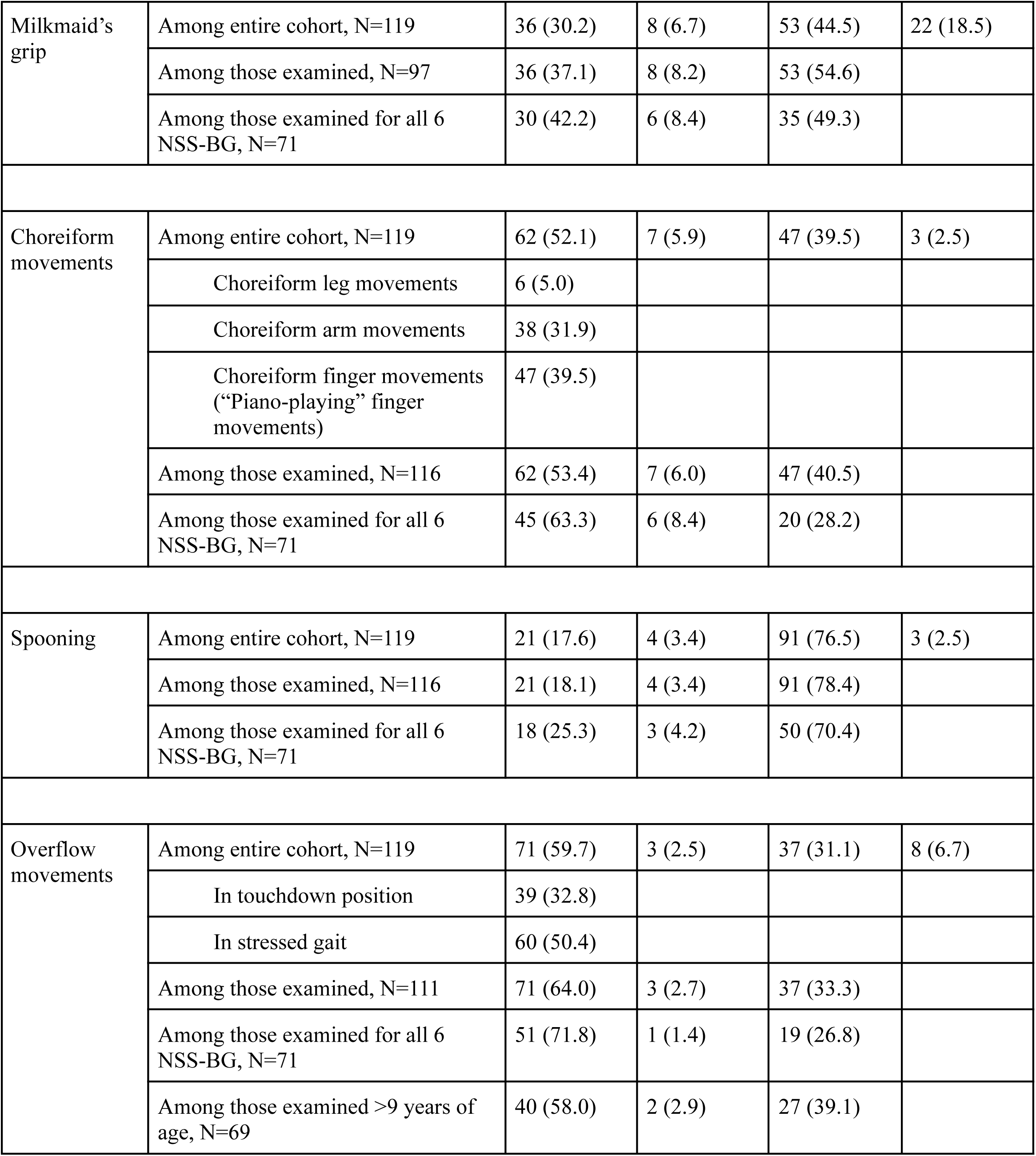
Prevalence of neurological soft signs reflective of basal ganglia dysfunction (NSS-BG) for 119 consecutive patients with pediatric acute-onset neuropsychiatric syndrome (PANS) at presentation.

All 119 patients were examined for at least one NSS-BG. Of the cohort, 79.8% (N=95/119) presented with at least one NSS-BG, and the mean number of NSS-BG was 2.1. Among those who had a complete neurological exam (N=71/119, 59.7%), meaning they were examined for all six NSS-BG, 91.5% (N=65/71) presented with at least one NSS-BG, and the mean number of NSS-BG was 2.6 (Figure 1).

**Figure 1.**
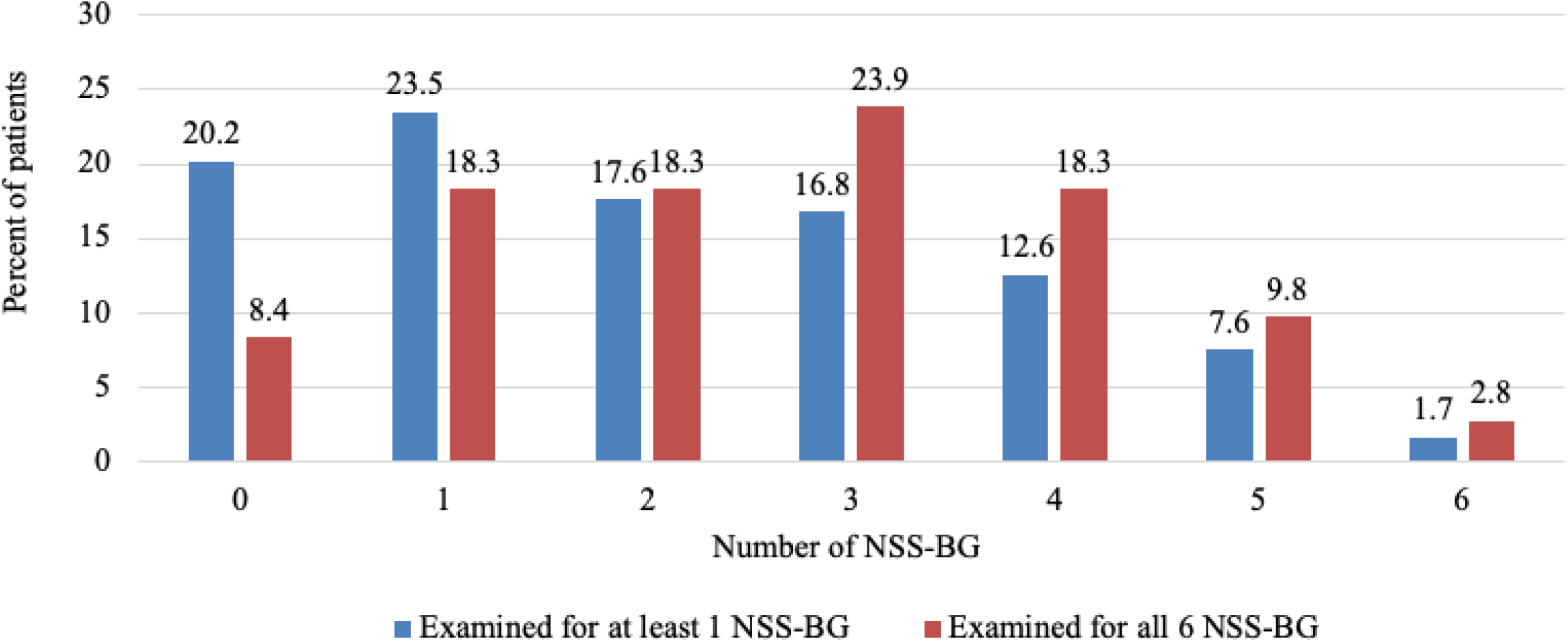
Number of neurological soft signs reflective of basal ganglia dysfunction (NSS-BG) in 119 consecutive patients with pediatric acute-onset neuropsychiatric syndrome (PANS) at presentation.

A bar graph illustrating the percentage of patients with each number of NSS-BG at clinical presentation of PANS. All 119 patients were examined for at least one NSS-BG. 71 patients were examined for all six NSS-BG. NSS-BG include glabellar tap reflex, abnormal tongue movements, milkmaid’s grip, choreiform movements, spooning, and overflow movements.

Compared with the 24 patients with 0 NSS-BG, the 95 patients with ≥1 NSS-BG and the 26 patients with ≥4 NSS-BG had more PANS symptoms (11.5 vs. 13.6 symptoms, p=0.04 and 11.5 vs. 15.1 symptoms, p=0.008) and shorter PANS duration (3.2 vs 1.9 years, p=0.03 and 3.2 vs 1.5 years, p=0.04). Additionally, compared with the patients with 0 NSS-BG who provided GIS (N=13/24), patients with ≥4 NSS-BG who provided GIS (N=22/26) reported worse global functioning of borderline significance (GIS: 56.0 vs 40.6, p=0.052). There was no significant difference in age or CBI (Table 4).

**Table 4.** Comparison of age, PANS duration, and disease severity at presentation between patients with and without neurological soft signs reflective of basal ganglia dysfunction (NSS-BG)

| | Patients with 0 NSS-BG (N=24) | Patients with $\geq 1$ NSS-BG (N=95) | Patients with $\geq 4$ NSS-BG (N=26) |
| --- | --- | --- | --- |
| Age (years) at the first clinic visit, mean (SD) | 10.4 (4.8) | 9.7 (3.2) | 9.8 (2.6) |
| PANS duration, mean (SD) | 3.2 (3.3) | 1.9 (2.5)** | 1.5 (2.5)** |
| Number of PANS symptoms, mean (SD) | 11.5 (4.2) | 13.6 (4.3)** | 15.1 (4.9)*** |
| Global Impairment Score (GIS), mean (SD) | 40.6 (26.7)<br>(N=19) | 43.8 (22.4)<br>(N=88) | 56.0 (22.6)*<br>(N=22) |
| Caregiver Burden Inventory (CBI), mean (SD) | 32.2 (18.4)<br>(N=13) | 32.1 (17.0)<br>(N=62) | 32.0 (17.2)<br>(N=17) |

\*p=0.052 (compared with patients 0 NSS-BG)
\*\*p<0.05 (compared with patients 0 NSS-BG)
\*\*\*p<0.01 (compared with patients 0 NSS-BG)

On Poisson regression, neither age at first visit (p=0.40) nor PANS duration (p=0.09) predicted the number of NSS-BG (models 1 and 2). Model 3 showed that the number of NSS-BG was significantly associated with the number of PANS symptoms at clinic presentation (1.049, 95% CI: 1.018-1.082, p=0.002). On linear regression, model 4 showed that the number of NSS-BG was significantly associated with Global Impairment Score (GIS) (2.857, 95% CI: 0.092-5.622, p=0.045).

There was no significant association found between overflow movements and age ≥9 years. There was also no significant association found between spooning and joint hypermobility.

## Discussion

This is the first study to describe the prevalence of neurological soft signs which may reflect basal ganglia dysfunction (NSS-BG) in patients with PANS at clinical presentation. We observed a high frequency of NSS-BG in patients with PANS who presented to the Stanford Children’s Immune Behavioral Health (IBH) Clinic, with more NSS-BG associated with greater symptomatology and impairment.

Previous studies have reported neurological findings in patients with PANS at the time of MRI^6^ as well as during evaluation at the NIH.^49^ As shown in Table 5, these studies are fairly consistent with the findings from this study. The slightly higher rates of NSS-BG in previous studies could be explained by selection bias: those patient cohorts are representative of more severe cases, as only a small fraction of PANS patients receive imaging or need specialized care from the NIH program.

**Table 5.**
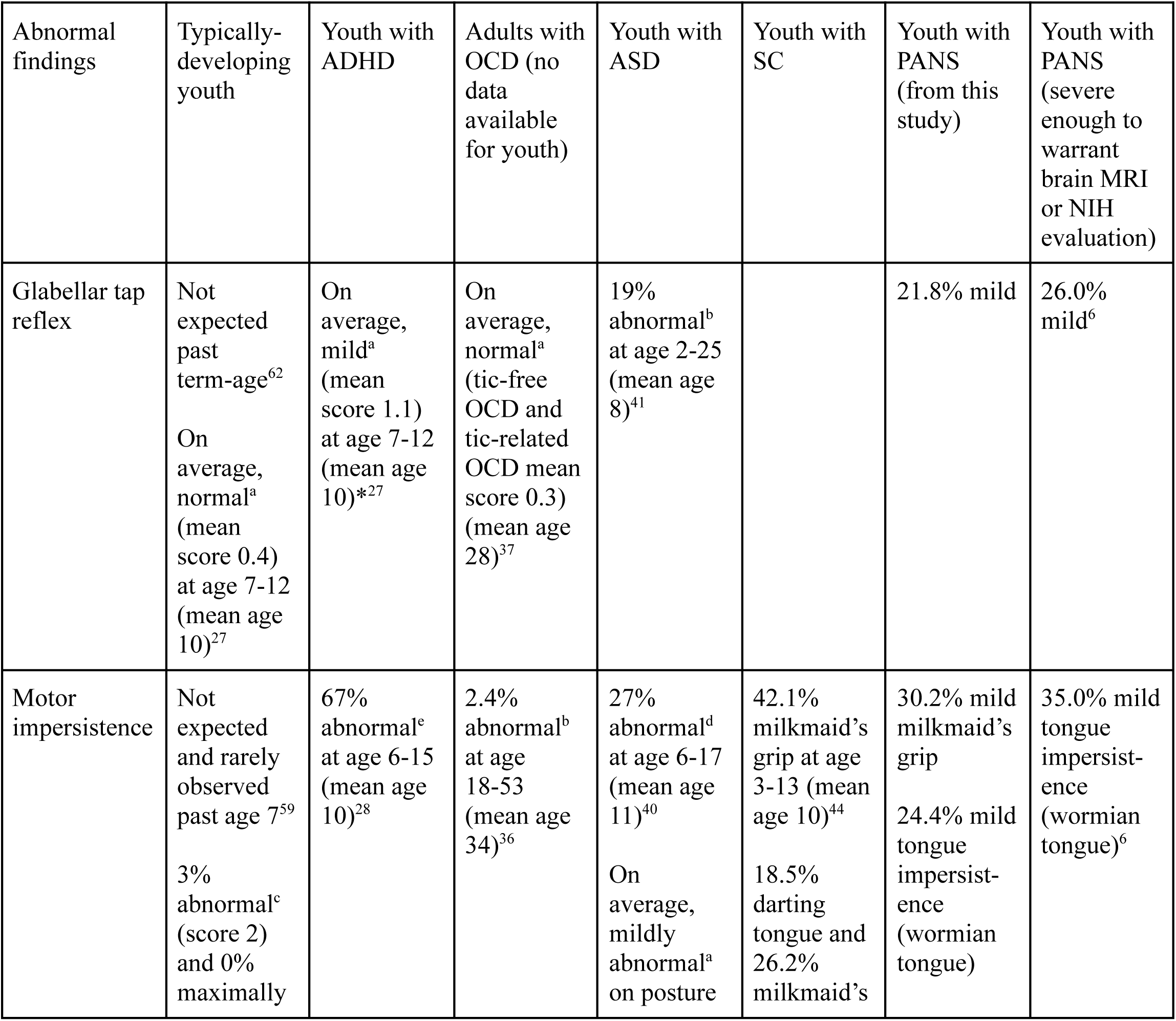
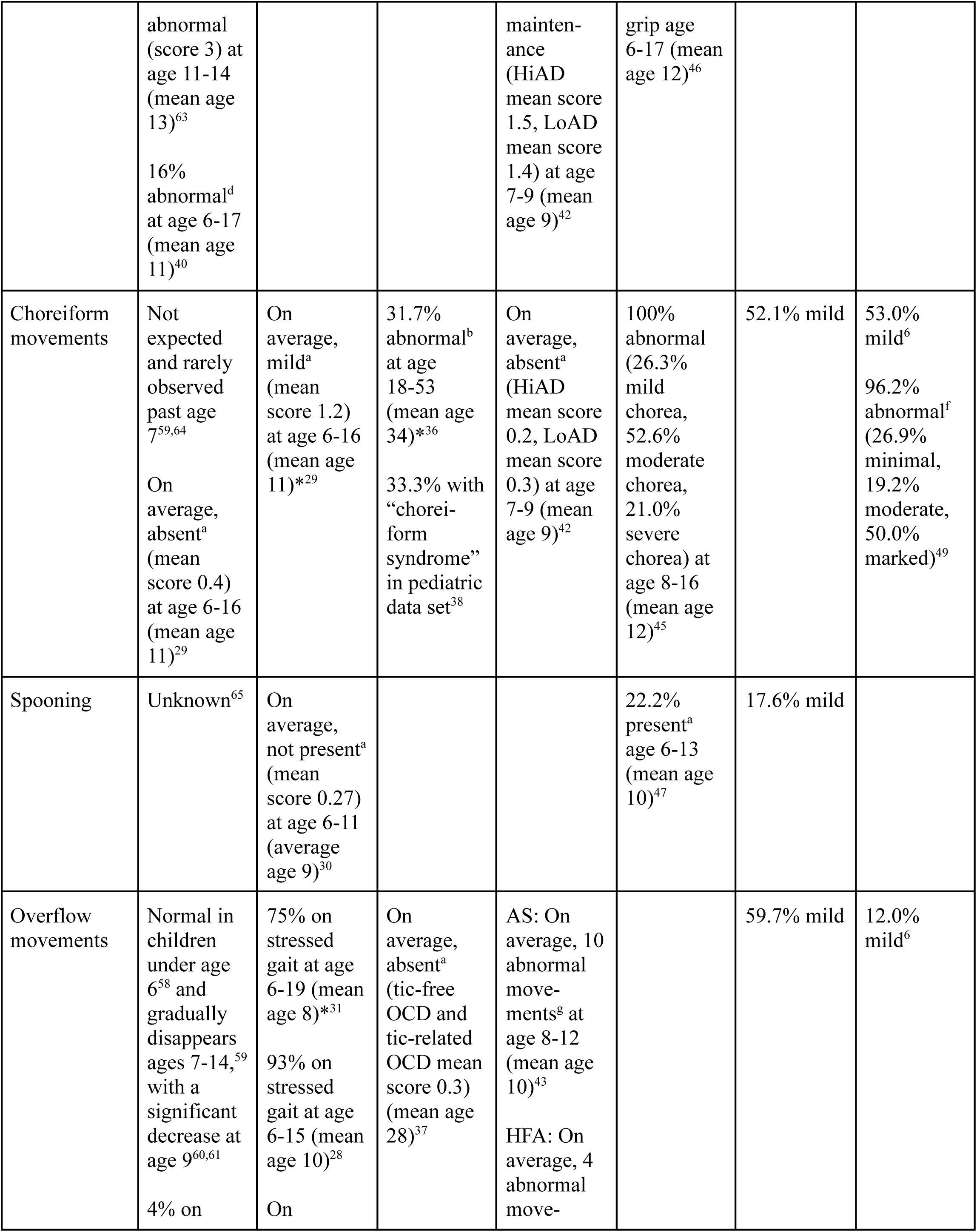
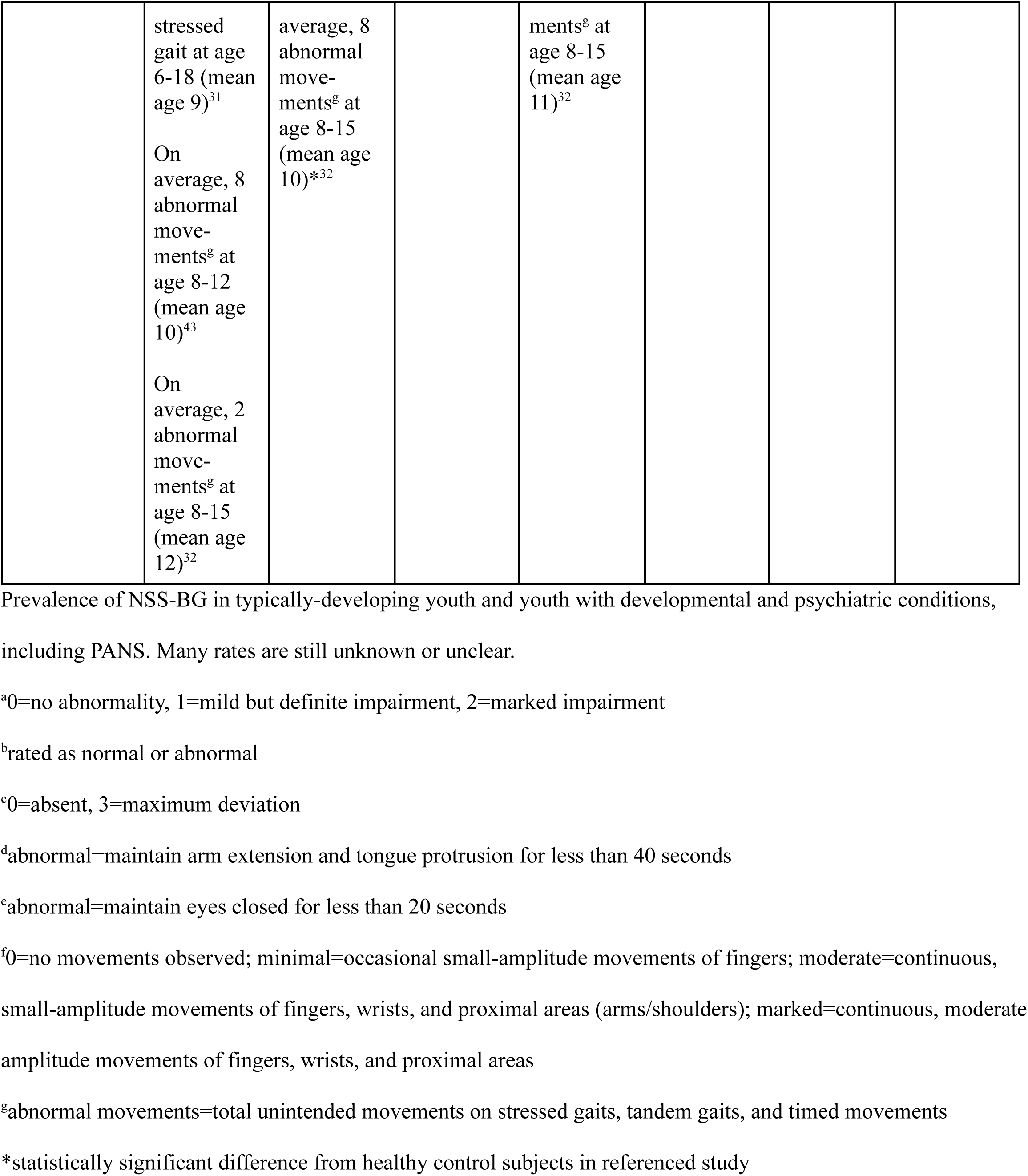
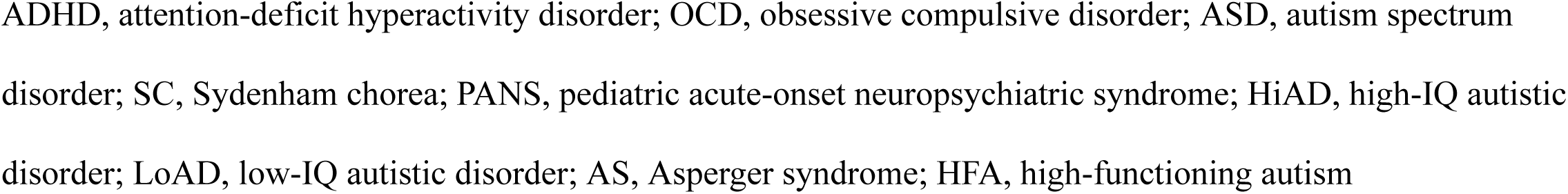
Rates of neurological soft signs which may reflect basal ganglia dysfunction (NSS-BG) in developmental and psychiatric populations.

Neurological soft signs are subtle exam findings thought to represent abnormalities in the development of neurologic circuits.^54^ These are contrasted with “hard neurological signs,” such as chorea and dystonia, which are often associated with structural and/or genetic basal ganglia abnormalities.^55^ However, the distinction on exam is not always clear, as choreiform movements and motor overflow may be mild forms of chorea and dystonia, respectively. This has important clinical implications. For example, in a child with acquired chorea in the absence of genetic findings, Sydenham chorea is the probable diagnosis. However, if the chorea is more subtle (“choreiform”), then the child is more likely to fall under the PANS category, assuming the other PANS criteria are met. Since there is often not a clear line between chorea and choreiform movements, research classification and enrollment in trials are challenging in borderline cases.

Based on a PubMed search (the formal strategy for which is included in eAppendix 2 of the Supplement), NSS-BG are variably seen in developmental and psychiatric conditions, including ADHD, OCD, autism, schizophrenia, and Sydenham chorea (Table 5). NSS-BG have been evaluated in adults with OCD,^33,36,37,56,57^ but there are no studies on NSS-BG in youth with OCD. A book chapter^38^ reported that 33% (n=18/54) of youth with OCD age 6-20 had “choreiform syndrome,” but this term was not defined.

Based on our literature review (Table 5), NSS-BG are typically not observed in school-age children, with the exception of overflow movements, which are normal in children under age 6^58^ and gradually disappears ages 7-14,^59^ with a significant decrease at age 9.^60,61^ Additionally, younger age was not associated with more NSS-BG in our study, further confirming that the unusually high prevalence of NSS-BG in our cohort is not attributable to early developmental stage.

Similar to what is reported in the literature,^60,61^ in our cohort, patients younger than or equal to age 9 had a higher frequency of overflow movements compared to patients older than age 9 (74% vs 58%). However, this difference was not statistically significant. Moreover, the rates of overflow movements in both the younger and older subset are notably high compared to the expected in typically-developing youth (Table 5), confirming the validity of evaluating this finding in patients with PANS, especially those above the age 9 threshold.

While there are >70 neurological soft signs and >160 exam maneuvers,^66^ the exam maneuvers used in this study were chosen because PANS is a suspected basal ganglia inflammatory disorder^3–9^ and these maneuvers are sensitive to basal ganglia dysfunction. However, the pathophysiology is likely more complex and other circuits may be involved. For example, in the glabellar tap sign, the basal ganglia and interconnected cortical structures have a defined role in the timing and reciprocity of blinking. Thus, persistent blinking may reflect basal ganglia dysfunction and/or frontal lobe dysfunction.^67,68^ Inability to maintain tongue protrusion and milkmaid’s grip are both manifestations of the motor impersistence seen in chorea. Tongue deviation could be due to dystonia but can also be seen in tongue weakness or structural abnormalities.^69,70^ However, in our patients with tongue deviation (N=10), there were no structural changes or tongue atrophy, and the tongue deviation improved as the flare resolved. Overflow movements are involuntary posturing in body regions outside of the area of intentional movement and may be secondary to pathology in the basal ganglia, cerebellum, and sensorimotor cortex.^71^ Given the preponderance of data that implicate basal ganglia involvement in PANS,^3–9^ our proposed set of NSS-BGG likely reflect abnormalities in circuits that involve the basal ganglia. It is also possible that pre-existing dysfunction in the basal ganglia predated and predisposed to PANS. Thus, it will be important to determine the relationship between NSS-BG and clinical state. If these NSS-BG resolve after patient recovery, this would contradict basal ganglia involvement being a static predisposing factor.

Some clinicians have theorized that spooning is a manifestation of joint hypermobility rather than dystonic posturing, given the high prevalence of hypermobility in patients with PANS.^72,73^ In our PANS cohort, there was no significant association found between spooning and joint hypermobility, suggesting it may be a relevant dystonic feature in PANS and should be evaluated. Nonetheless, future studies should define whether the prevalence of spooning is associated with other NSS-BG. More broadly, the ability to distinguish joint and muscle symptoms from subtle abnormal neurological findings should be further explored in this patient population.

In our study, a higher number of NSS-BG was associated with more PANS symptoms and a greater impairment score. In the Zheng et al.^6^ imaging study, patients with choreiform movements at the time of MRI had more diffuse brain abnormalities. In synthesis, patients with PANS who present with NSS-BG may have not only greater inflammation but also greater disease severity. This suggests that NSS-BG may be helpful in the diagnosis and clinical monitoring of PANS, which currently has no reliable biomarker. Given that most youth with PANS undergo relapses and remissions (84%),^74^ during which they alternate between symptomatic flares (often following infection) and improvements toward baseline after several months, determining patient clinical state is crucial to managing symptoms and treatment. Most patients (N=105, 88.2%) were in a flare during the physical examination at presentation, so the study may not have been powered for assessing the relationship between NSS-BG and disease state. Further study is needed to understand the evolution of NSS-BG during the course of disease.

Some NSS-BG may be helpful for differentiating between developmental and psychiatric conditions. For example, choreiform movements are typically absent in youth with autism^42^ but are commonly present in youth with PANS,^6,49^ ADHD,^29^ OCD,^38^ and Sydenham chorea.^45^ Thus, if a patient diagnosed with autism exhibits choreiform movements, further diagnostic workup for other pediatric developmental and psychiatric conditions may be warranted. Since many NSS-BG observed in our patients with PANS are also frequently observed in patients with ADHD, OCD, autism, and Sydenham chorea (Table 5), the presence of a specific NSS-BG may not distinguish PANS from these other conditions. However, these conditions could have unique NSS-BG profiles which could aid in diagnosis. This possibility underscores the significance of completing and analyzing neurological exams for these pediatric populations. Moreover, given that neurological soft signs (NSS) are often general markers of psychopathology, they may be relevant to determining the neuropsychiatric status of the patient, regardless of diagnosis.

There are some limitations to this study. Although the neurological exam was standardized, variability in exam methods and findings may have occurred between clinicians, and some data was missing. We were unable to routinely conduct a comprehensive neurological exam including NSS-BG at every first visit, and exams were sometimes performed during remission or the resolution phase of the flare. In fact, 14 of the 119 patients (11.8%) had their NSS-BG evaluated for the first time when they were not in a flare state. Four of those 14 patients (28.6%) were not examined until the second or third visit. A challenge with this patient population is that patients who are doing poorly from the behavior/mental health standpoint may not be able to cooperate with neurological exams but may exhibit high levels of basal ganglia dysfunction (21.8% (N=26/119) of comprehensive neurological exams including NSS-BG had to be deferred to future visits). Thus, we may have underestimated the true prevalence of NSS-BG in patients with PANS. Lastly, the prevalence of NSS-BG in healthy youth and in other disorders including OCD, ADHD, and autism, and Sydenham chorea is not well established, so it is unclear if presence of NSS-BG allow us to distinguish PANS from other pediatric conditions.

## Conclusions

Children and young adults presenting with PANS commonly have multiple neurological soft signs which may reflect basal ganglia dysfunction (NSS-BG) on physical examination, with more NSS-BG reflecting greater impairment as measured by PANS symptoms and global levels of functioning. While the diagnosis of PANS is based primarily on history, these findings suggest that targeted neurological exams may help support diagnosis. Furthermore, the high frequency of NSS-BG in PANS provides further evidence that the pathogenic mechanisms of the condition may involve the basal ganglia, which could be the target of psychotherapeutic medications and rehabilitation. As these exams are often not routinely performed by neurologists or pediatricians, we advocate for their use in evaluating youth for immune-mediated neuropsychiatric deterioration.

## Supporting information

Supplement_eAppendix1

Supplement_eAppendix2

## Data Availability Statement

The raw data supporting the conclusions of this article will be made available by the authors, without undue reservation.

## Ethics Statement

This study was reviewed and approved by the Stanford Institutional Review Board. Written informed consent to participate in this study was obtained from all adult participants and from the parent/legal guardian/next of kin of all participants aged less than 18 years.

## Author Contributions

JZ and JF contributed to the conception and design of the study. JG and JF created the data fields and definitions of neurological examination findings for a separate project that was published (Zheng et al.).^6^ JF and JG trained JZ and BV, who abstracted neurological soft signs from medical records. This data was audited by JG and JF using the Zheng et al. dataset. MT, MS, PT, and YX classified patients as PANS. BF, JF, MM, and PT completed neurological exams. JZ, JG, and LT developed and performed the statistical analysis. CS and JZ developed and executed the search strategy. JZ, BV, JG, JW, and JF wrote the manuscript. All authors contributed to manuscript review, read, and approved the submitted version.

## Acknowledgments

We would like to thank Susan Swedo, MD (developmental pediatrician, National Institutes of Health), Arnold Gold, MD (general neurologist, Columbia University), and Terence Sanger, MD, PhD (movement disorder specialist, neurology, University of California, Irvine) for teaching of the neurological soft signs which may reflect basal ganglia dysfunction and answering follow-up questions regarding patients and this manuscript.

We would also like to thank current and former members of our research staff, our IBH Clinical Team, the Stanford PANS Basic Science Team, our collaborating physicians, and NAPPA. We are especially grateful for our patients and families who understand treatment limitations and continue to lend their time and cooperation to research participation.

