## Supplement_eAppendix1 for "Prevalence of Neurological Soft Signs at Presentation in Pediatric Acute-Onset Neuropsychiatric Syndrome"

eAppendix 1. Criteria for Diagnosis of PANS[1]

1. Abrupt, dramatic onset of obsessive-compulsive disorder or severely restricted food intake
2. Concurrent presence of additional neuropsychiatric symptoms, with similarly severe and acute onset, from at least two of the following seven categories:
    a. Anxiety
    b. Emotional lability and/or depression
    c. Irritability, aggression and/or severely oppositional behaviors
    d. Behavioral (developmental) regression
    e. Deterioration in school performance
    f. Sensory or motor abnormalities
    g. Somatic signs and symptoms, including sleep disturbances, enuresis or urinary frequency
3. Symptoms are not better explained by a known neurologic or medical disorder, such as Sydenham chorea, systemic lupus erythematosus, Tourette disorder or others.

eReference.

1. Swedo SE, Leckman JF, Rose NR. From Research Subgroup to Clinical Syndrome: Modifying the PANDAS Criteria to Describe PANS (Pediatric Acute-onset Neuropsychiatric Syndrome). *Pediatrics & Therapeutics.* 2012;2(2):1-8. doi:10.4172/2161-0665.1000113
