## Supplement_eAppendix2 for "Prevalence of Neurological Soft Signs at Presentation in Pediatric Acute-Onset Neuropsychiatric Syndrome"

eAppendix 2. PubMed Search Strategy

1. ("Autism Spectrum Disorder"[Mesh] OR "Obsessive-Compulsive Disorder"[Mesh] OR "obsessive compulsive" [tw] OR austis* [tw])
2. ("dyskinesias" [mesh] OR "chorea" [mesh] OR "neurologic examination" [mesh] OR choreiform [tw] OR "soft sign*" [tw] OR glabellar [tw] OR spooning [tw] OR overflow [tw] OR milkmaid [tw] OR tongue [tw] OR (movement [tw] AND impersist* [tw]))
3. (infan* [tw] OR newborn* [tw] OR "new-born" [tw] OR "new borns" [tw] OR perinat* [tw] OR neonat* [tw] OR baby [tw] OR baby* [tw] OR babies [tw] OR toddler* [tw] OR minors [tw] OR child [tw] OR child* [tw] OR children* [tw] OR schoolchild* [tw] OR "school child" [tw] OR "school children" [tw] OR adolescen* [tw] OR juvenil* [tw] OR youth* [tw] OR teen* [tw] OR "under age" [tw] OR pubescen* [tw] OR prepubesc* [tw] OR pediatric* [tw] OR paediatric* [tw] OR "Nursery school" [tw] OR kindergar* [tw] OR "primary school" [tw] OR "secondary school" [tw] OR "elementary school" [tw] OR "high school" [tw] OR "primary schools" [tw] OR "secondary schools" [tw] OR "elementary school" [tw] OR "high schools" [tw] OR highschool* [tw] OR "infant" [mesh] OR "Child"[mesh] OR "Adolescent"[mesh] OR "Minors" [mesh] OR "Puberty" [mesh] OR "Pediatrics" [mesh] OR pediatrics) AND english [lang]
4. 1 AND 2 AND 3
